# Genetic hallmarks of patients with congenital portosystemic shunts and portopulmonary hypertension: protocol for a multicentre exploratory study

**DOI:** 10.64898/2026.10.27.26366476

**Authors:** Isabelle Schepens, Julie Wacker, Frédéric Pont, Frank van Steenbeek, Raphael Joye, Maurice Beghetti, Periklis Makrythanasis, Valérie A. McLin

**Affiliations:** Department of Paediatrics, Gynaecology and Obstetrics, University of Geneva, Geneva, Switzerland; Paediatric Cardiology Unit, Department of Paediatrics, Gynaecology and Obstetrics, University of Geneva, Geneva, Switzerland; Communication in Science, Geneva, Switzerland; Department Clinical Sciences, Faculty of Veterinary Medicine, Utrecht University, Utecht, The Netherlands; Paediatric Cardiology Unit, Division of Paediatric Specialties, Department of Woman, Child and Adolescent Medicine, Geneva University Hospitals and Faculty of Medicine, University of Geneva, Geneva Switzerland; Laboratory of Medical Genetics, Medical School, ‘Aghia Sophia’ Children’s Hospital, National and Kapodistrian University of Athens, 115 27 Athens, Greece; Biomedical Research Foundation of the Academy of Athens, 115 27 Athens, Greece; Department of Genetic Medicine and Development, Medical School, University of Geneva, CH-1211 Geneva 4, Switzerland; Swiss Pediatric Liver Center, Gastroenterology, Hepatology and Pediatric Nutrition Unit, Department of Pediatrics, Gynecology and Obstetrics, University of Geneva, Geneva, Switzerland

**Keywords:** Congenital Portosystemic Shunt, Pulmonary Arterial Hypertension, Whole Genome Sequencing, Genetic Predisposition to Disease, Pedigree

## Abstract

**Introduction:** Congenital portosystemic shunts (CPSS) are rare vascular malformations, with an estimated prevalence of 1/30’000 live births, that cause portal blood to partly bypass the liver. CPSS frequently co-occur with congenital heart disease and other malformations, suggesting a shared developmental or genetic basis. Severe acquired clinical manifestations are present in a large proportion of patients with CPSS. Portopulmonary hypertension (PoPH), the most life-threatening, has been reported in as many as 7–14% of CPSS patients.

Mounting evidence indicates that PoPH can arise early in life, particularly in children with CPSS, yet paediatric PoPH remains understudied. This project aims to identify genetic variants associated with PoPH in CPSS patients through whole-genomes, focusing primarily on known pathogenic genes for pulmonary arterial hypertension (PAH), congenital heart disease (CHD), Hereditary Haemorrhagic Telangiectasia (HHT), SNPs previously associated with PoPH in adults, and ultimately novel genomic variants. The overarching goal of the study is to improve personalised risk assessment and inform clinical decisions regarding elective shunt closure in individuals potentially at-risk of developing PoPH.

**Methods and analysis:** The aim is to perform an international multi-centre exploratory genome analysis of patients with CPSS and PoPH identified through the International Registry of Congenital Portosystemic Shunts (IRCPSS). Whole-genome sequencing will be utilized to analyse both coding and non-coding regions in a predominantly paediatric cohort. Targeted analysis of established pathogenic genes associated with PAH, CHD, HHT, and PoPH will be complemented by a genome-wide approach to identify novel, potentially pathogenic variants, with variant classification assessed according to American College of Medical Genetics (ACMG) criteria. Family-based analyses, including trio analyses and segregation studies, will enable the identification of dominant, recessive, and de novo variants, facilitating a comprehensive evaluation of variant inheritance pattern and pathogenic potential.

**Ethics and dissemination:** This study has been approved by the CCER (Geneva cantonal ethics committee for research) and poses minimal risk to participants, limited to blood sampling performed as part of routine clinical care for patients. Genetic data will be securely stored and analysed in compliance with GDPR. Clinically significant findings will be communicated through recruiting physicians, in accordance with participant preference regarding incidental findings. Study results will be disseminated through open-access peer-reviewed publications and scientific conferences to advance understanding and management of PoPH in individuals with CPSS and potentially other chronic liver diseases associated with portosystemic bypass.

**ClinicalTrials.gov:** NCT07314814

**CCER Ethical approval:** 2024-01698

**STRENGTHS AND LIMITATIONS OF THIS STUDY:**

- Systematic analysis of established gene panels for PAH, CHD and HHT, to diagnose referenced genetic disorders.
- The trio-based whole-genome analysis design allows for the detection of novel potentially pathogenic variants and genes.
- The rarity of CPSS limits recruitment to an expected sample size of ~30 trios, both a realistic target for a rare disease study and appropriate family-based analyses.
- Leverages the International Registry of Congenital Portosystemic Shunts as a resource for correlating genetic and clinical data

## Introduction

Congenital Portosystemic Shunts (CPSS) are rare venous malformations that lead to partial or complete diversion of portal blood away from the liver, leading to direct communication between porto-mesenteric blood and the systemic circulation. CPSS result from impaired vascular remodelling during early development. The estimated prevalence of umbilico-porto-venous shunt – thought to be the precursors of CPSS – in foetuses has been reported to range between 1/478– 1/595 pregnancies in tertiary care, and from 1/6110 to 1/9357 pregnancies in primary care settings [1–3]. The incidence of CPSS at birth has been estimated at 1/30’000–1/50’000 based on galactosemia screening [4]. However, the true incidence among live-born children is likely underestimated because asymptomatic patients remain undetected. 50–70% of individuals with CPSS eventually develop one or more associated clinical manifestations [5–7]. Some may arise early in life, including neonatal high-output cardiac failure, cholestasis, or primary liver tumours, or may develop later in the course of the disease.

Among complications, pulmonary arterial hypertension (PAH) stands out as the most life-threatening. It is reported in as many as 7–14% of patients [4,8]. When PAH occurs in individuals with CPSS, it is referred to as PoPH (portopulmonary hypertension). PoPH is a recognised indication for CPSS closure to mitigate the risk of death [8]. Therefore, understanding which patients are at risk of developing PoPH is essential. PoPH in the setting of CPSS is a very rare disease, which makes anticipatory management challenging.

Pathogenic or likely pathogenic variants in at least 17 genes have been implicated in PAH. Some of these variants are associated with an early disease onset. However, the penetrance of disease-causing variants is incomplete, and disease onset may vary considerably according to several factors, including sex, particularly after puberty. The International Consortium for Genetic Studies in PAH has published recommendations for genetic counselling across the spectrum of PAH subtypes [9]. Gaining a better understanding of the genetic factors underlying PAH in patients with CPSS may help identify individuals at increased risk of developing PoPH and inform anticipatory management.

The present study leverages the unique population enrolled in the International Registry of Congenital Portosystemic Shunts (IRCPSS) to overcome this obstacle and identify genetic hallmarks candidates associated with susceptibility to PoPH in CPSS patients. These findings may ultimately yield insights that are generalizable to the broader population of patients with cirrhotic and non-cirrhotic portal hypertension. The IRCPSS was created to characterize the clinical phenotype of patients with CPSS (CE 2018-00413) [10]. To date, it includes longitudinal comprehensive clinical data of over 450 subjects from 27 centres worldwide, including cardiopulmonary assessment, essential for PoPH documentation. It serves as a valuable resource to identify subjects likely to contribute to the present study on the genetic characteristics of PoPH in the CPSS population. The underlying study rationale is that patients presenting early in life with severe PoPH in the setting of CPSS likely harbour a unique genetic background predisposing them to early and severe disease, above and beyond the consequences of portosystemic bypass alone, a recognised trigger of pulmonary vascular disease.

The objective of the study described in this protocol is to identify genetic variants associated with the clinical development of PoPH in individuals with CPSS. The study will focus on established gene panels for PAH, for congenital heart disease (CHD) [9] as well as for hereditary haemorrhagic telangiectasia (HHT), with which significant common denominators are expected because of shared vascular development pathways. Association with CPSS has been observed both for CHD [11–13] and HHT [14–17].

*De novo* variants have emerged as an important class of genetic factors underlying rare diseases, especially severe early-onset conditions [18–22], as highly damaging mutations reduce reproductive fitness and are therefore rarely inherited [23,24]. Accordingly, the present study also adopts a broader non-targeted approach, based on whole-genome analysis, to identify potential novel variants associated with CPSS-PoPH. Whole-genome sequencing capacity enables comprehensive analysis of the entire human genome, both coding and non-coding regions, with single-nucleotide resolution. A crucial element of the study design is the inclusion of comparative genetic analyses of the child and both parents (trio-based) to identify not only *de novo* variants but also biallelic variants in trans in the case of recessive variants.

## Methods and Analysis

The study will involve approximately 30 families, each comprising a proband with a prior diagnosis of CPSS-PoPH and both biological parents. Each family will be analysed independently, and no formal aggregate statistical analysis is planned. This approach is consistent with genetic studies of rare diseases aimed at identifying pathogenic variants exhibiting dominant, recessive, or *de novo* inheritance patterns. The target sample size of at least 30 families is based on our group’s expertise in the genetics of rare diseases [25], and reflects a realistic recruitment goal for this exceptionally rare condition. This cohort is expected to enable the identification of potentially pathogenic variants and candidate genes that can be further investigated and validated in future studies.

### Project population

The study population is recruited via the network of experts collaborating in the IRCPSS (CE 2018-00413) [10]. At the time of writing, more than 63 participants (out of more than 450) are categorised as CPSS and pulmonary hypertension. Of these, 5 were excluded because PH was diagnosed during the neonatal period, leaving 58 with features of PoPH. The cardiopulmonary data of these 58 potential participants will be thoroughly reviewed to determine eligibility before patients are invited to participate in this study. Considering that participants in the IRCPSS and their legal representatives have already demonstrated their interest in participating in a research project, it is estimated that a sufficient proportion of patients and their families will consent to participate in the present study.

The study population is composed of children and adult patients, their siblings, and their parents. There is no age limit for the recruitment of patients, siblings, or parents. The inclusion and exclusion criteria are conceived to be compatible with the study’s methodological requirements, while maximizing patient inclusion.

Inclusion Criteria:

- Patient is enrolled in the IRCPSS with a diagnosis of PoPH.
- Trio consisting of a patient with PoPH and both biological parents (only complete trios are eligible).
- Sibling of an enrolled patient.
- The patient and both parents agree to provide biological samples (blood) and sign the informed consent.
- Siblings or their legal representatives agree to provide biological samples (blood) and sign the informed consent.
- Exclusion criteria:
  ∘ Offspring–parent trios resulting from adoption or donor-assisted reproductive technology.
  ∘ Sibling pairs with at least 1 non-shared parent.
  ∘ Secondary PSS (i.e. acquired rather than congenital).

### Project presentation, consent request, and compensation

Participants will be recruited internationally from centres participating in the IRCPSS. An online webinar will be organised to ensure that all recruiting physicians are trained and compliant with the ICH E6 Good Clinical Practice guidelines. The session will also provide detailed instructions on the study’s scope, protocol procedures, the information sheet, informed consent documentation, and the eCRF.

Documentation will be provided for project presentation and consent forms formulated in local language and in an age-adapted format for potential participants. The recruiting physician or investigator will have an informative discussion with the families about the project, covering the scope of the study, the procedures involved, the expected duration, the potential risks and benefits and any discomfort. Each participant will also be informed that the participation in the study is voluntary and that they may withdraw from the study at any time without affecting subsequent medical assistance and treatment. Participants will be informed that their medical records may be examined by authorised individuals other than their treating physician for the purposes of the study and controls from the ethical authorities. Any questions will be answered. Informed consent will be obtained from the participants or their legal guardians as appropriate.

In view of future studies on CPSS, participants will be given the option to sign additional independent consents to store and reuse residual biological samples and data generated for the present study, and to collect additional blood samples for biobanking.

Participants will not receive any financial compensation for participation in the study. Reimbursement of travel expenses will be provided. Overnight accommodation may be offered, as needed.

### Sample collection

Trios and siblings will be invited for a single visit (preferably during a regular clinical visit) for blood sampling.

The volume of collected EDTA blood will be based on HUG (Hôpitaux Universitaires de Genève) standards (minimal volume for adult: 3 mL, children >2y.o. 1 mL). Twice the volume will be collected if the participant accepts additional sample collection for biobanking. Blood draw will be performed under topical anaesthesia for children. For subjects younger than 2 years old, a mouth swab will be proposed to allow minimal genetic analysis in case the age of 2 years cannot be reached before the end of the recruitment/sample collection period.

### Sample preparation and whole-genome sequencing

Sample preparation and whole-genome sequencing (WGS) will be performed either locally or in Geneva. For WGS performed locally, the raw data (FASTQ) will be transferred to the analysis team in Geneva on a secured platform to ensure analytical consistency and reduce systematic analytical biases.

#### Methods at sponsor site

##### a) DNA extraction

Blood samples will be transported to the HUG genomics and molecular diagnostics laboratory (DIAGMOL). A local coded identifier will be attributed to the samples upon reception, as per the DIAGMOL standard operating procedure. This local code will be paired with the IRCPSS coded identifier. The DIAGMOL code will be kept for the duration of sample processing and will be used for genomic data labelling. This code will be recorded in the eCRF for DIAGMOL-IRCPSS identifier matching. During dataset processing, the code correspondence key may be accessed by the coordinating investigator or the co-investigator in charge of genetic analyses to facilitate the identification and management of offspring–parent trio datasets.

The DIAGMOL will prepare, quantify, and aliquot genomic DNA for sequencing. Genomic DNA will be extracted from fresh blood within 5 days of sample collection. In case this time interval cannot be met, samples will be frozen until processing. DNA samples will be pre-aliquoted according to Genome Center standard operating procedure and placed at a registered position in a barcoded plate. The plate will then be transported to the Genomic platform of the Genome Center (Geneva). Leftover DNA will be stored in the local biobank of the DIAGMOL for an unlimited period or treated according to participant consent. This DNA may be used later in the present study to confirm variants by the accredited DIAGMOL clinical laboratory using Sanger sequencing or Multiplex Ligation-dependent Probe Amplification (MLPA).

In case the participant consents to the collection of additional blood, the additional sample will also be managed by the DIAGMOL laboratory as described above. Each sample will be attributed a local identifier and will be stored in the local DIAGMOL Biobank.

##### b) Sequencing

Whole-genome sequencing will be performed at the sequencing platform of the Genome Center (Geneva). Upon receipt, samples will be stored at −20°C until further processing for controls and analysis. A quality control (fragment analyser, concentration check) will be performed before including samples in the WGS pipeline.

Samples will be sequenced within one month of receipt without prior gDNA amplification, as DNA sequencing for variant screening requires high-quality and deep-coverage (>30X) sequencing. Samples will be processed until the target of 30X depth is reached. Once sequencing output has passed quality control, the raw data FASTQ files will be uploaded on the Swiss-based Saphetor VarSome platform for analysis.

Depending on the nature of the variants identified, participants may be called for complementary medical examination for a phenotyping visit lasting 1-2 hours.

#### Methods at collaborator sites

Material and data transfer agreements will be established between the centres before any sharing. Participating centres will share material and data to integrate the workflow at different steps: frozen blood, genomic DNA, raw genomic data, and clinical information. Most centres have expressed a preference for transferring genomic datasets rather than blood or DNA samples. Centres will be advised to perform genomic DNA preparation and sequencing in a specialised accredited facility for genomics (ISO 15189 certified where possible). When appropriate, samples will be shipped to the relevant facility in Geneva as needed. Raw genomic data can be directly uploaded on the Swiss-based Saphetor platform. All samples or data will be received coded.

### Genomic data analysis

Analysis of WGS data will follow standard procedures up to variant annotation, where ACMG 2015 guidelines will be used.

Analytical genomic analysis pipelines will be selected at the time of analysis to incorporate the latest methodological developments and maximise analytical output. The criteria for selection of pipelines are:

1. The current state of the art in bioinformatics tools
2. The hardware requirements for the tools (especially if AI is used) and storage.
3. The time required for data analysis
4. Budgetary constraints.

As most of the subjects will likely not have previously undergone genetic testing, the proband will be analysed first for diagnostic purposes. Diagnostic gene panels will include genes for the known clinical presentation of the patient, and for similar or associated disorders (PAH, CHD, HHT, PoPH). If a pathogenic variant is identified, family members will be tested to further characterise the pathogenicity of the variant and its inheritance pattern. Cases with a pathogenic variant will be referred to the clinical genetics diagnostic laboratory at HUG for independent confirmation in a medically accredited laboratory (DIAGMOL in Geneva). Confirmed diagnosis will be communicated to the recruiting physician and to the participant as per consent. If no genetic diagnosis is identified, the family will be included in the main study cohort for the identification of novel genetic variants.

To facilitate meaningful variant interpretation both for coding [26] and non-coding [27] variants the relevant ACMG guidelines will be used. These guidelines take into account parameters such as:

1) type of amino-acid change, functional element alteration, 2) absence or low population allele frequency in the GnomAD Browser database, 3) various deleteriousness prediction scores such as CADD, 4) publications demonstrating the functional effect of the variant at the time of the analysis (PubMed, NCBI), 5) sequence conservation (PhastCons, PhyloP, GERP++), 6) gene constraint using the probability of loss-of-function intolerance (pLI score, GnomAD), 7) variant interpretation resources (ENCODE, 1000 Genome project, GnomAD) and others. Additionally, newer tools harnessing AI, such as AlphaFold, will be used to increase the prediction accuracy of the pathogenicity of genomic variants.

Dominant and recessive variants will be sought, adapting the strategy depending on which individual – parent or sibling – is/are affected within a family, in case another individual than the patient is also affected within the family. The genetic workflow for variants identification is provided in Figure 1, and the study principles in Table 1.

**Table 1.** Study principles for variants identification.

| Step | 1*- Panels | 2*- Dominant analysis |  |  | 3*- Recessive analysis |
| --- | --- | --- | --- | --- | --- |
| **Affected individuals** | Independent of how many family members are affected | Patient only | Patient + 1 sibling | Patient + 1 parent | Patient, parents, and other affected members |
| **Type of analysis** | Targeted analysis | Non-targeted analysis | Non-targeted analysis | Non-targeted analysis | Non-targeted analysis |
|  | Diagnostic panel analysis | Search for *de novo* variants | Search for *de novo* variants | Search for vertical transmission | Search for recessive genes |
| **Genome analyses** | Patient genome | Trios: patient + parents; genomes (genes and regulatory elements) | Family: patient + parents + siblings; genomes (genes and regulatory elements) | Family: patient + parents + siblings; genomes (genes and regulatory elements) | Trios or Family; genomes (genes and regulatory elements) |
| **Principle** | Analysis focused on specific gene panels associated with similar diseases or related pathologies | Analysis workflow to identify *de novo* variants | Principles of *de novo* variants identification apply; analysis will be performed independently for each affected sibling. | The process will be adapted on a case-by-case basis depending on family segregation | Analysis workflow to identify homozygote or compound heterozygote variants |
| **Outcome** | Pathogenic variant in a known gene | For each trio/family, the outcome is a list of variants with estimated pathogenicity. |  |  |  |
| **Family testing** | Pathogenic variant identified in subject genome; diagnostic testing will be pursued for other family members | In case a likely pathogenic variant is identified in (at least) one individual, its presence or absence in additional family members will be assessed and linked to potential phenotypic features. |  |  |  |

**Figure 1.**
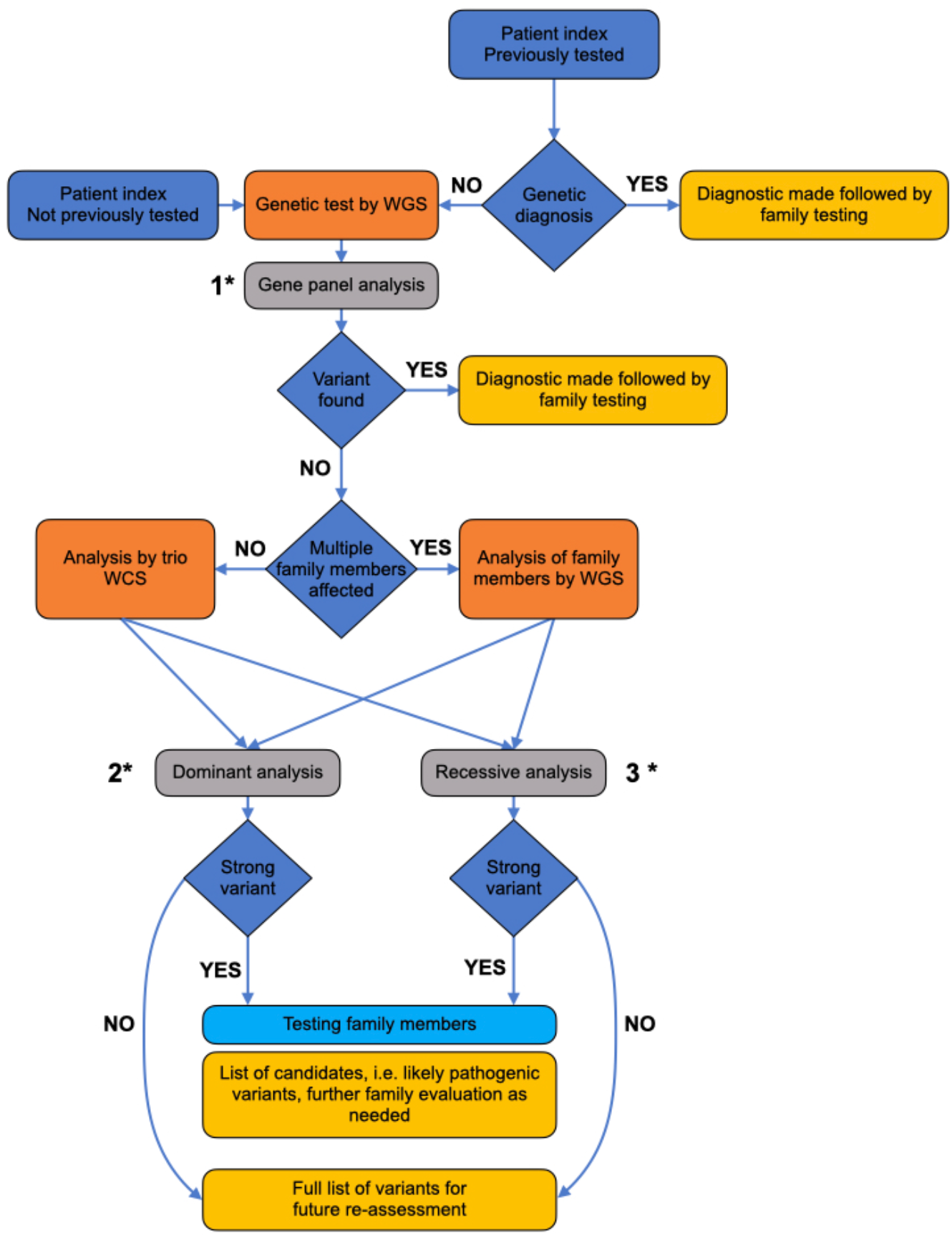
Genetic workflow for variants identification. summarises the workflow of the present study, including participants that were or were not previously assessed genetically during their management.

#### Dominant analysis

The goal of this analysis is to identify *de novo* or inherited dominant variants. In the most common scenario, where only one patient in the family is affected, the analysis will focus on the patient and their parents (trio analysis), targeting *de novo* variants. *De novo* variants arise due to DNA replication errors during spermatogenesis or oogenesis and are thus absent in the germline of the parents. The human germline mutation rate per generation is 1-2 × 10^−8^ [28] resulting in 40– 80 *de novo* single-nucleotide variants per individual genome. Given the fact that the coding part of human genome is ~1% of the total, usually only a single variant is located in a protein-coding region [29].

For families where multiple members are symptomatic, the analysis will be adjusted based on family structure as follows:

- If a sibling is symptomatic: a separate *de novo* trio analysis will be conducted for each sibling. Before evaluating individual variants, a comparison will be performed to identify variants shared among both siblings – which indicates parental germline mosaicism. These shared variants will be prioritised for further evaluation.
- If a parent is symptomatic: the analysis cannot rely on *de novo* filtering, and will therefore focus on vertically transmitted variants, shared between the affected parent and the child. Only variants in protein-coding exons will be evaluated to reduce the number of candidates to be analysed for two reasons: 1) because this approach yields a much larger number of candidate variants compared with *de novo* analysis and 2) because grand-parents are not part of the current study. Depending on the total number of patients/families analysed, burden analyses will be conducted to identify genes, genomic elements and gene pathways that contribute to the disease.

#### Recessive analysis

Analysis to detect recessive causes of CPSS-PoPH will be performed. Using trio data, the phase of biallelic variants will readily be detected. Only variants in trans located in coding regions and known functional elements will be included, as the total number of variants is expected to be significant, akin to that observed in cases with an affected parent.

The analysis will be adapted as follows if more than one family member is affected:

- If a sibling is affected: only genes containing the same variants in both patients will be considered.
- IfIf a parent is affected, the search for recessive genes will proceed as usual, but the candidate variants will be examined on a case-by-case basis, since vertical transmission of rare recessive diseases is extremely unlikely outside of consanguineous families or isolated populations.

### Study endpoints

The primary endpoint of the present study is a list of genetic variants in coding and noncoding genomic regions, encompassing:

1. pathogenic variants in genes included in established clinical diagnostic panels for similar or associated conditions (PAH, HTT, CHD)
2. potentially pathogenic variants located in novel genes or previously unexplored genomic regions,

The secondary endpoint is the comprehensive list of all other genetic variants identified in patients with CPSS-PoPH.

### Biases of the study

The main sources of bias in this study arise during the bioinformatic analysis and interpretation of genetic data. The uneven knowledge between coding and non-coding regions introduces an interpretation bias: assessing the significance of the variants in non-coding regions is more challenging than assessing that of variants in better characterised coding regions. Furthermore, there is a selection bias in favour of a younger population, because inclusion criteria require both parents for trio analysis, something easier to achieve in paediatrics.

Finally, an ascertainment bias may be present because more severe and clinically recognized cases are likely overrepresented in the IRCPSS. However, this selection effect also increases the feasibility and potential yield of genetic discovery in an exceptionally rare disease.

## Ethics and dissemination

### Ethical approval

This study has received approval from the Geneva cantonal ethics commission for research (CCER), approval 2024-01698, and will be conducted in accordance with the Declaration of Helsinki, the principles of Good Clinical Practice, the Human Research Act (HRA), the Human Research Ordinance (HRO), and all other applicable regulations. Any substantial protocol amendments will be submitted to the Ethics Committee for approval before implementation.

### Benefits

Participants are not expected to receive direct clinical benefit, except for patients for whom a diagnosis is obtained. However, the knowledge generated may contribute to improved risk stratification and management of children and adults with CPSS who are at risk of developing PoPH in the future.

Clinically relevant incidental findings that fall within established reporting guidelines and that are confirmed by an accredited diagnostic laboratory will be returned to participants if they have consented to receive such information. Incidental findings unrelated to CPSS or PoPH, including misattributed parentage, will not be investigated or disclosed.

### Data management and confidentiality

Data will be collected and stored in a secure, pseudonymised REDCap database hosted on institutional servers with controlled access, audit trails, and regular backups. Genomic data will be generated and stored in accordance with institutional and national data protection standards. Only coded data will be transferred to the centralised server for genomic sequences analysis of the study. Raw sequencing data and derived analysis files will be stored on secure servers for the duration permitted by ethics approval and participant consent.

### End of project procedures

Upon study completion or early termination, the Ethics Committee will be informed. Biological samples will be retained or destroyed according to participant consent and institutional procedures. Genomic data and key analysis files will be archived on secure servers for long-term preservation, as required by Swiss law.

### Dissemination

The findings of this study will be disseminated through peer-reviewed publications, international scientific conferences, and presentations to the IRCPSS network. Summaries of study results will be made available to participating families upon request. No identifiable data will be included in any dissemination material.

## Data Availability

All data produced in the present study are available upon reasonable request to the authors

## Acknowledgements

The authors gratefully acknowledge Timothée Brütsch from Communication in Science for his valuable support in the preparation of this manuscript.

## Footnotes

### Contributions

∘ Conceptualization: VML, PM, FvS, IS
∘ Data curation: JW, RJ, PM, IS, VM
∘ Formal analysis: N/A
∘ Funding acquisition: VML (Lead), IS, MB
∘ *Investigation: N/A*
∘ Methodology: PM (Lead), IS, VM
∘ Project administration: VML, IS
∘ Resources: VM
∘ *Supervision: N/A*
∘ *Validation: N/A*
∘ *Visualization: N/A*
∘ Writing – original draft: IS, VML, PM, FP
∘ Writing – review & editing: JW, MB, RJ, FvS

### Patient and Public Involvement

There was no patient or public involvement in the preparation of the present study.

